# Human-AI Collaboration in Large Language Model-Assisted Brain MRI Differential Diagnosis: A Usability Study

**DOI:** 10.1101/2024.02.05.24302099

**Authors:** Su Hwan Kim, Severin Schramm, Cornelius Berberich, Enrike Rosenkranz, Lena Schmitzer, Kerem Serguen, Christopher Klenk, Nicolas Lenhart, Claus Zimmer, Benedikt Wiestler, Dennis M. Hedderich

## Abstract

**Background:** Prior studies have shown the potential of large language models (LLMs) to support in differential diagnosis in radiology. However, the interaction of human users with LLMs in this context has not been evaluated.

**Purpose:** To investigate the impact of human-LLM collaboration on accuracy and efficiency of brain MRI differential diagnosis.

**Methods:** In this retrospective study, twenty brain MRI cases with a challenging but definitive diagnosis were selected and randomized into two groups. Six inexperienced radiology residents were instructed to determine the three most likely differential diagnoses for each of these cases via conventional internet search or utilizing an LLM-based search engine (© Perplexity AI, powered by GPT-4). Accuracy of suggested differential diagnoses was analyzed using the chi-square test and Mann-Whitney U test. Interpretation times were analyzed using the student’s t-test. Benefits and challenges in human-LLM interaction were derived from observations and participant feedback.

**Results:** LLM-assisted brain MRI differential diagnosis yielded superior accuracy (38/59 [LLM-assisted] vs 25/59 [conventional] correct diagnoses, p = 0.03). No difference in interpretation time (8.12 +/- 3.22 min [LLM-assisted] vs 7.96 +/- 2.65 min [conventional], p = 0.76) or level of confidence (median of 2.5 [LLM-assisted] vs 3.0 [conventional], p = 0.96) was observed. Several challenges related to human errors and technical limitations were identified.

**Conclusion:** Human-LLM collaboration has the potential to improve brain MRI differential diagnosis. Yet, several challenges must be addressed to ensure effective adoption and user acceptance.

## Introduction

Radiological differential diagnosis plays a crucial role in clinical care, profoundly influencing diagnostic and therapeutic decisions. Accurate determination of relevant differential diagnoses from image findings demands highly specialized knowledge of anatomy and pathophysiology along with proficiency in recognizing visual patterns and synthesizing comprehensive clinical information.

Recent studies suggest the emerging potential of large language models (LLMs) to execute radiological differential diagnosis based on case presentations (1–5). These studies compared the diagnostic suggestions of an LLM to expert assessments or confirmed diagnoses. Yet, the intricate interactions between human users and LLM systems in this context have not been explored.

Previous literature reveals the critical impact of human-AI interaction on diagnostic performance in radiology (6–8). One study employing an AI-based mammogram classification system demonstrated that inexperienced and experienced readers alike are susceptible to automation bias, which describes the inclination of human users to adhere to incorrect recommendations from automated decision-making systems (6). Similarly, incorrect AI results were shown to negatively impact radiologist performance in lung cancer detection based on chest radiography (7). Yet another study highlighted the significance of establishing effective human-AI collaboration protocols in AI-assisted knee MRI reading (8). Analogously, elements of human-AI collaboration could affect the outcomes of LLM-assisted differential diagnosis. In real-world practice, it is plausible that radiologists or radiology residents would use LLMs as an adjunct tool to support diagnostic reasoning rather than for fully autonomous differential diagnosis (9). Under these circumstances, the human medical professional assumes a pivotal position in contextualizing the available clinical and visual information, formulating the prompt, critically reviewing the LLM response, and conducting further research to eventually derive a conclusion. Particularly considering the well-known propensity of LLM systems to generate factually incorrect information (so-called hallucinations) (10,11), a comprehensive evaluation of how users realistically interact with these systems is imperative. Therefore, this study aimed to investigate the impact of human-LLM collaboration on accuracy and efficiency of brain MRI differential diagnosis.

## Methods

Ethical approval was waived by the institutional review board.

### Study Sample

Six radiology residents with less than 6 months of experience in neuroradiology were recruited from the local departments of radiology and neuroradiology as participants of the usability study and randomized into two groups. All participants provided informed consent. A total of twenty brain MRI exams obtained between 01/01/2016 and 12/31/2023 were selected from the local imaging database and randomized into two sets (Figure 1). In each brain MRI scan, the image finding in question was denoted by an arrow. Selection criteria for participants and MRI scans are shown in Table 1.

**Table 1:**
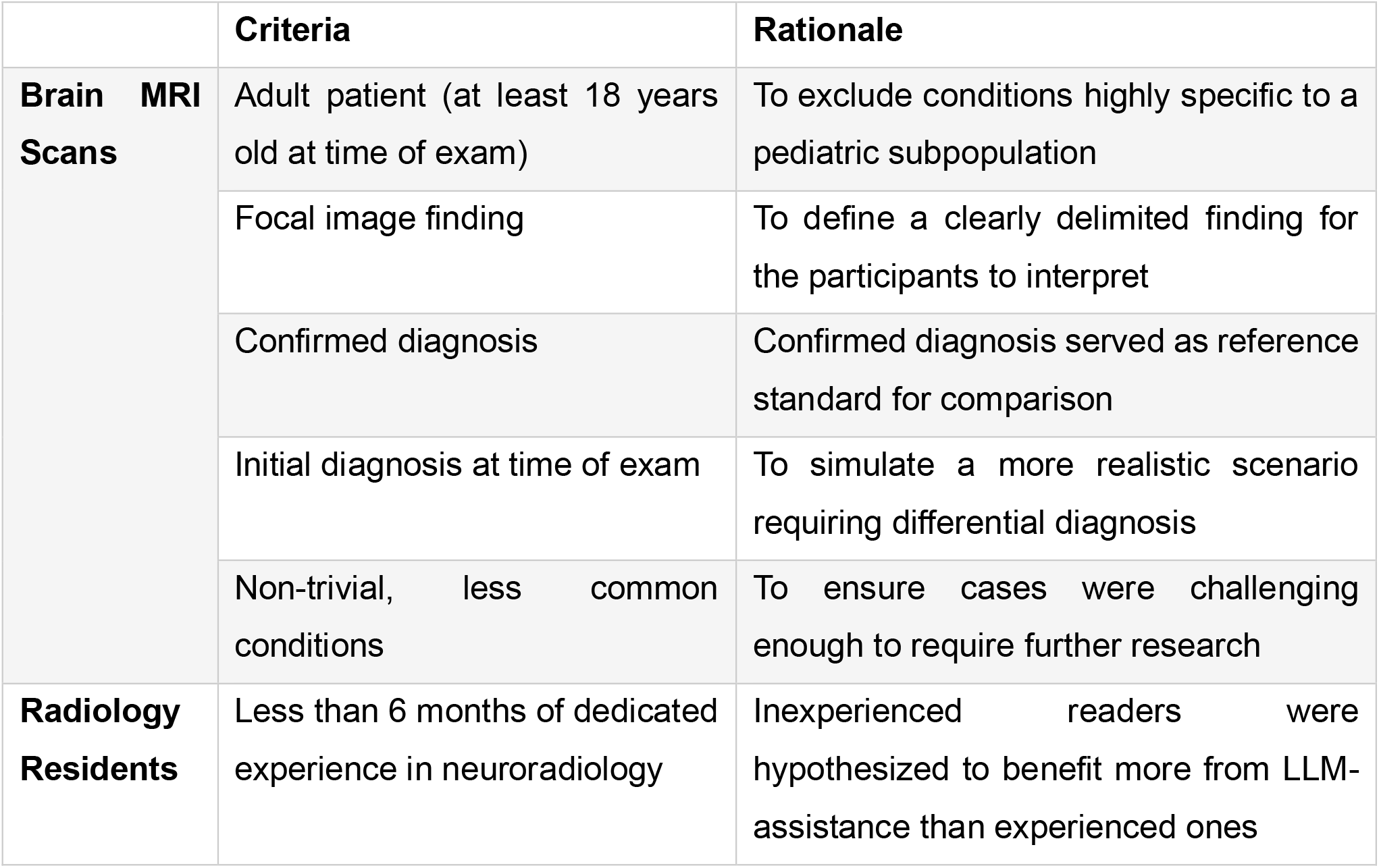
Inclusion Criteria.

|  | Criteria | Rationale |
| --- | --- | --- |
| **Brain MRI Scans** | Adult patient (at least 18 years old at time of exam) | To exclude conditions highly specific to a pediatric subpopulation |
|  | Focal image finding | To define a clearly delimited finding for the participants to interpret |
|  | Confirmed diagnosis | Confirmed diagnosis served as reference standard for comparison |
|  | Initial diagnosis at time of exam | To simulate a more realistic scenario requiring differential diagnosis |
|  | Non-trivial, less common conditions | To ensure cases were challenging enough to require further research |
| **Radiology Residents** | Less than 6 months of dedicated experience in neuroradiology | Inexperienced readers were hypothesized to benefit more from LLM-assistance than experienced ones |

**Figure 1:**
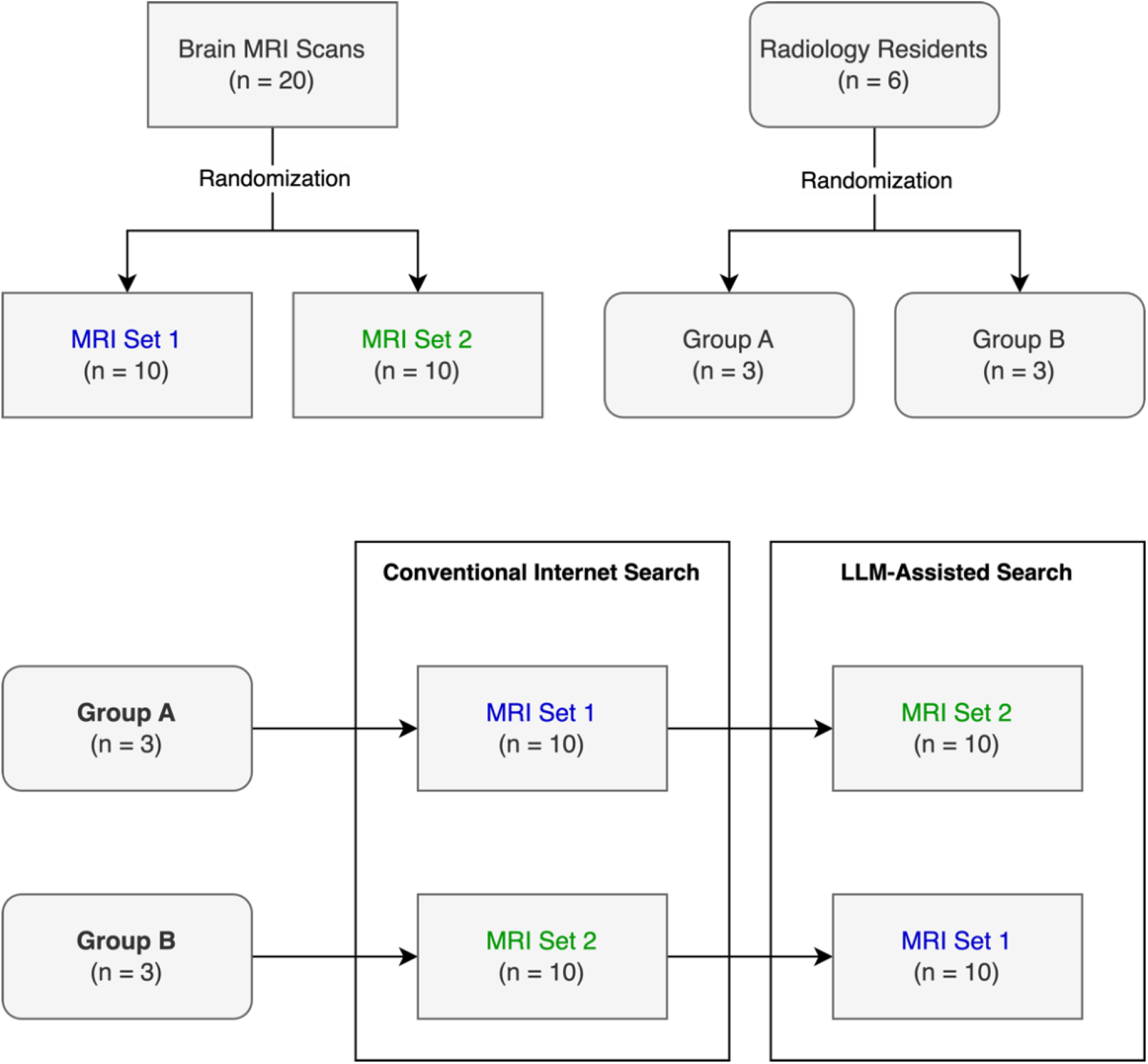
Study Design.

### Large Language Model (LLM) and Chatbot Interface

PerplexityAI (© Perplexity AI Inc., San Francisco, USA) was chosen as chatbot interface, given its ability to access real-time web content and to indicate its sources of information. GPT-4 (Generative Pre-trained Transformer 4), OpenAI’s most up-to-date LLM, was selected as the model powering the search queries.

### Study Design

Initially, the LLM system was introduced to participants in a 10–15-minute training session to ensure familiarity with its operation and functionality. During this training, participants were presented several sample prompts and explored the tool using a selection of sample brain MRI cases. Then, each participant was tasked to determine the most likely differential diagnoses of the annotated image finding in the brain MRI scans via conventional internet search for half (n = 10) and via LLM-assisted search (© PerplexityAI) for the other half (n = 10) (Figure 1). Cases were excluded from the analysis if participants were familiar with the case, or the image finding was not recognized despite the annotation. Importantly, participants performing LLM-assisted search were allowed to conduct additional conventional internet search to validate LLM suggestions. Participants were allowed to submit up to three differential diagnoses, ranked by relevance. Alongside the MRI scan, participants received demographic and clinical details of the patient case. All prompts were phrased in English. Interpretation times were measured using a time tracking software (Toggl Track, © Toggl OÜ, Talinn, Estonia). Level of confidence was recorded for each case on a Likert scale from 1-5 (1: very low confidence, 5: very high confidence). During the usability sessions, notes of relevant observations and comments were taken by SHK and SS. Before and after the sessions, participants completed a questionnaire to evaluate the LLM-assisted search workflow and capture concerns, benefits, and limitations.

### Analysis

To determine accuracy of differential diagnoses, two different scoring systems were applied. In the first approach, participant responses were classified as “correct” if the correct diagnosis was included among the submitted differential diagnoses and “incorrect” if it was not (binary scoring system). In the second approach, participant responses were assigned a score from 0 – 3, depending on the rank of the correct diagnosis within the response (numeric scoring system) (Table 2). Inferential statistics were performed using the chi-square test for binary scores and the Mann-Whitney U test for numeric scores and level of confidence. Interpretation times were analyzed using the student’s t-test. The level of significance was set at p < 0.05.

**Table 2:** Numeric scoring system for evaluating differential diagnoses.

| Score | Condition |
| --- | --- |
| 3 | Correct diagnosis ranked first |
| 2 | Correct diagnosis ranked second |
| 1 | Correct diagnosis ranked third |
| 0 | Correct diagnosis was not included in the response |

Likert-scale questions of the questionnaires were analyzed by descriptive statistics. Qualitative data from observation notes, comments and questionnaire responses were summarized in tables categorizing data by relevant topics. Data manipulation, analysis and visualization were performed using Python (version 3.9.7).

## Results

Two responses were excluded from the analysis due to the participant’s familiarity with the MRI scan or inability to recognize the annotated image finding. An overview of selected clinical cases is provided in Supplement 1.

### Quantitative Findings

LLM-assisted brain MRI differential diagnosis yielded superior accuracy, as evaluated by both binary (38/59 [LLM-assisted] vs 25/59 [conventional] correct diagnoses, p = 0.03) and numeric scoring approach (median score of 2 [LLM-assisted] vs 0 [conventional], p = 0.04) (Figure 2). No difference in interpretation time (8.12 +/- 3.22 min [LLM-assisted] vs 7.96 +/- 2.65 min [conventional], p = 0.76) was observed (Figure 3). Similarly, the level of confidence did not differ significantly (median of 2.5 [LLM-assisted] vs 3.0 [conventional], p = 0.96). A screenshot of a sample LLM response is shown in Figure 4. Questionnaire results revealed a moderately positive evaluation of the LLM-assisted workflow. Participants showed a slight tendency in favor of using the LLM tool in clinical practice (median: 4). Quality of LLM responses were rated rather positively (median: 4). The LLM system was easily adopted into the diagnostic workflow by most participants (median: 4). The overall experience of the LLM-assisted search workflow was mixed (median: 3.5) (Figure 5).

**Figure 2:**
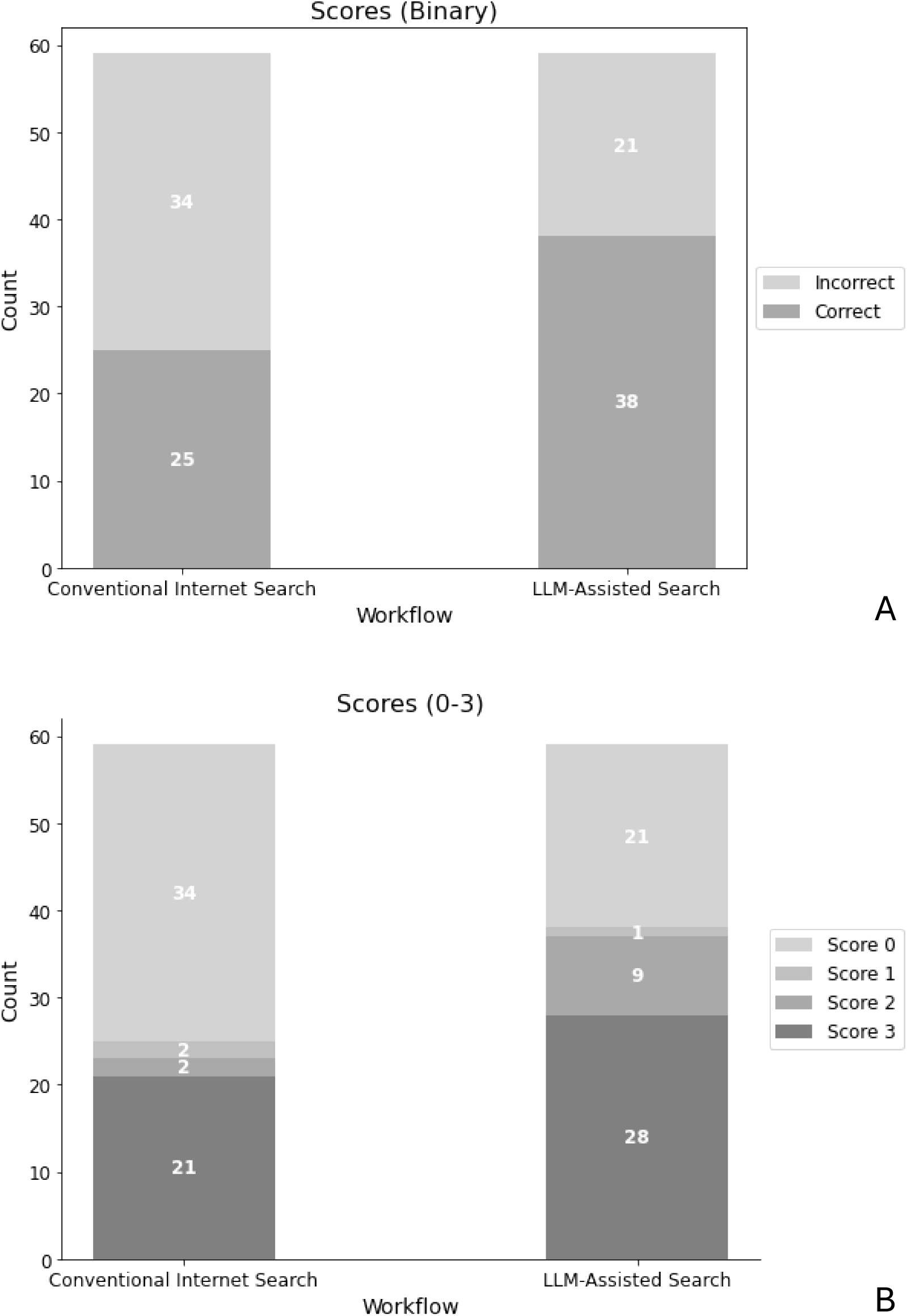
Accuracy of differential diagnoses by workflow. A: Binary scoring system. Participant responses were classified as either correct or incorrect. LLM-assisted workflow yielded superior scores (p = 0.03). B: Numeric scoring system. A participant’s response was assigned a score between 0 and 3, depending on the rank of the correct diagnosis within the response (3: correct diagnosis ranked first, 0: correct diagnosis not included in response). LLM-assisted workflow yielded superior scores (p = 0.04).

**Figure 3:**
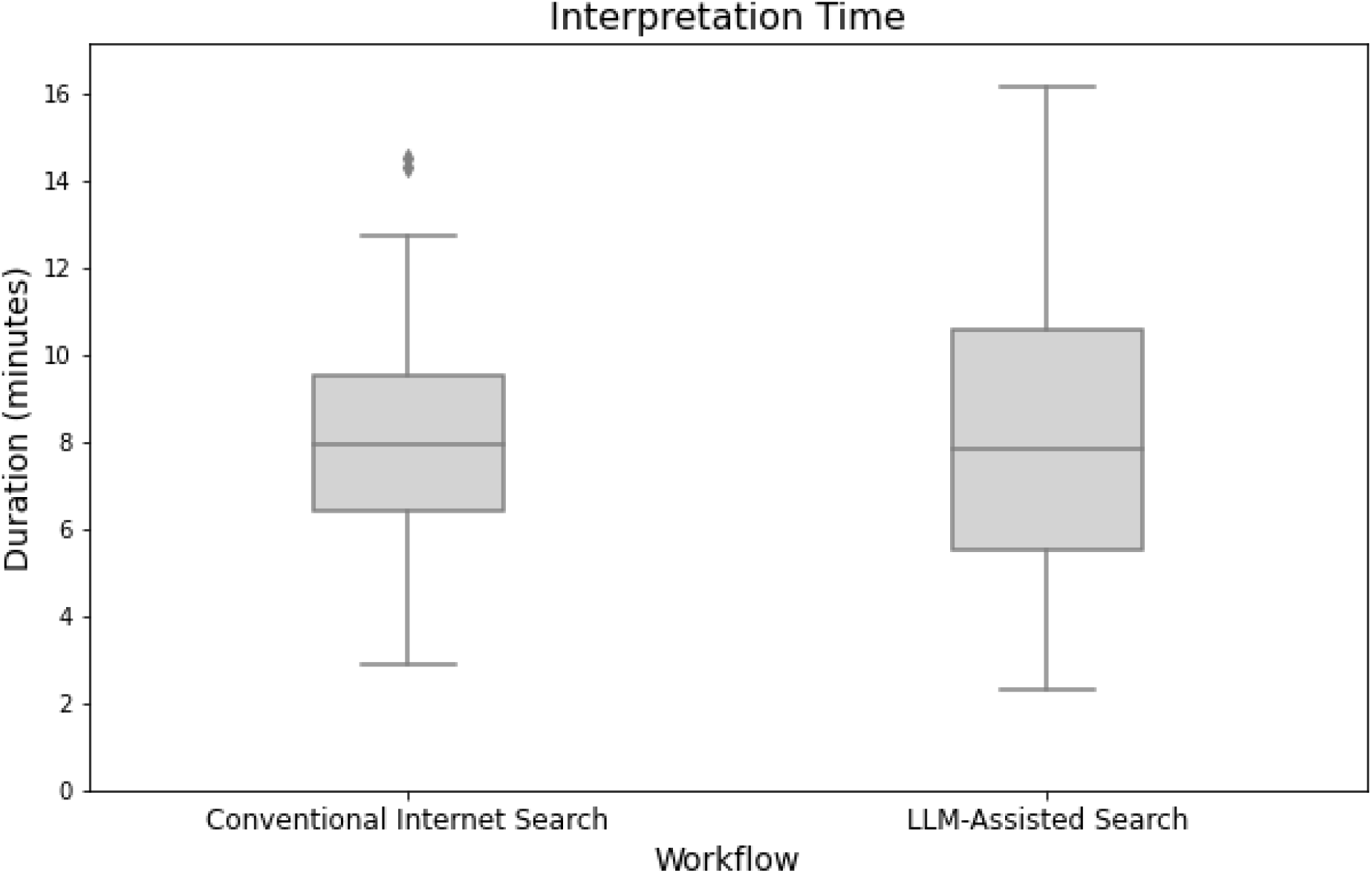
Interpretation time. Interpretation times did not exhibit any statistically significant difference (p = 0.76).

**Figure 4:**
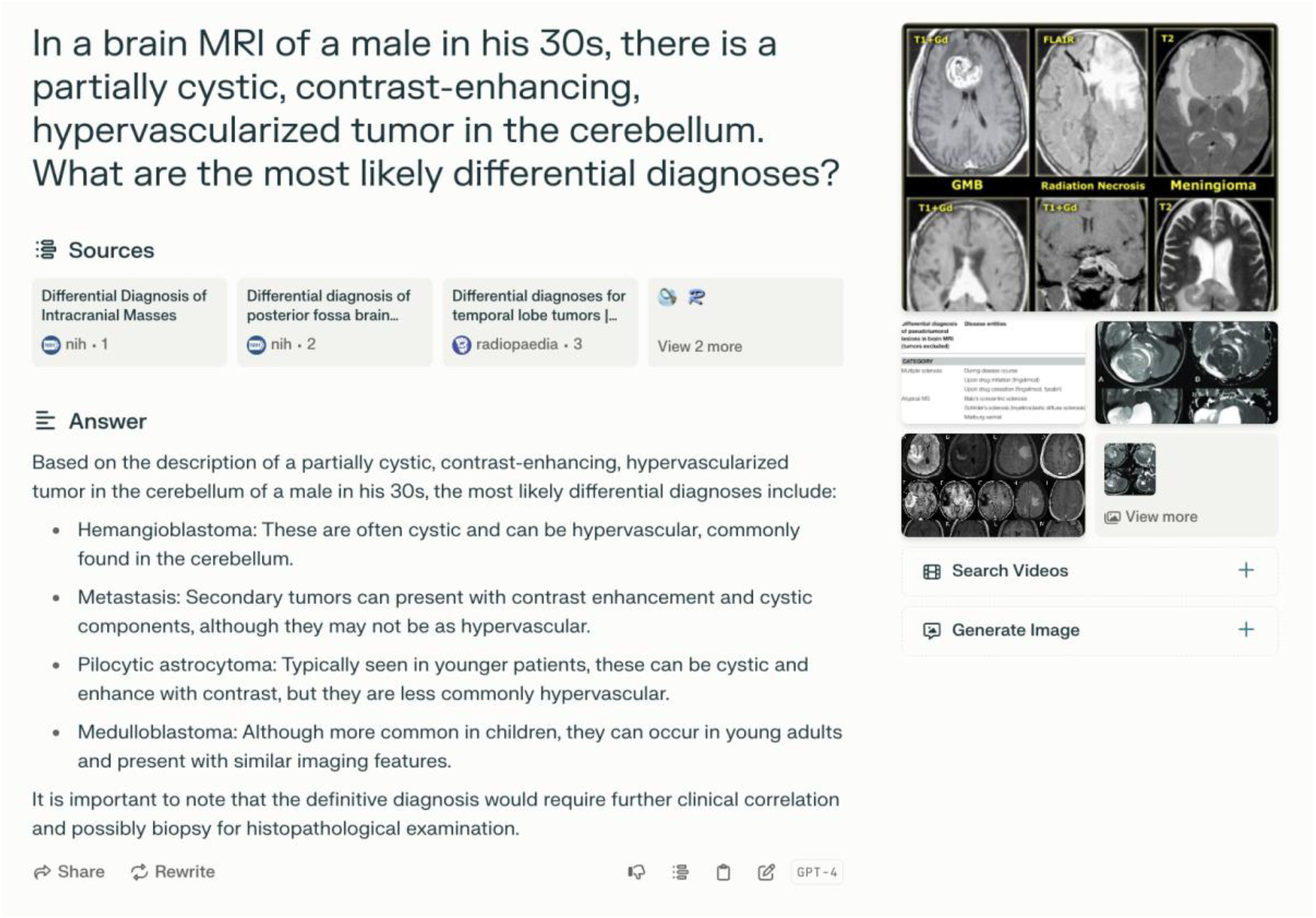
Screenshot of a sample LLM query (PerplexityAI). The correct diagnosis sought in this case was hemangioblastoma.

**Figure 5:**
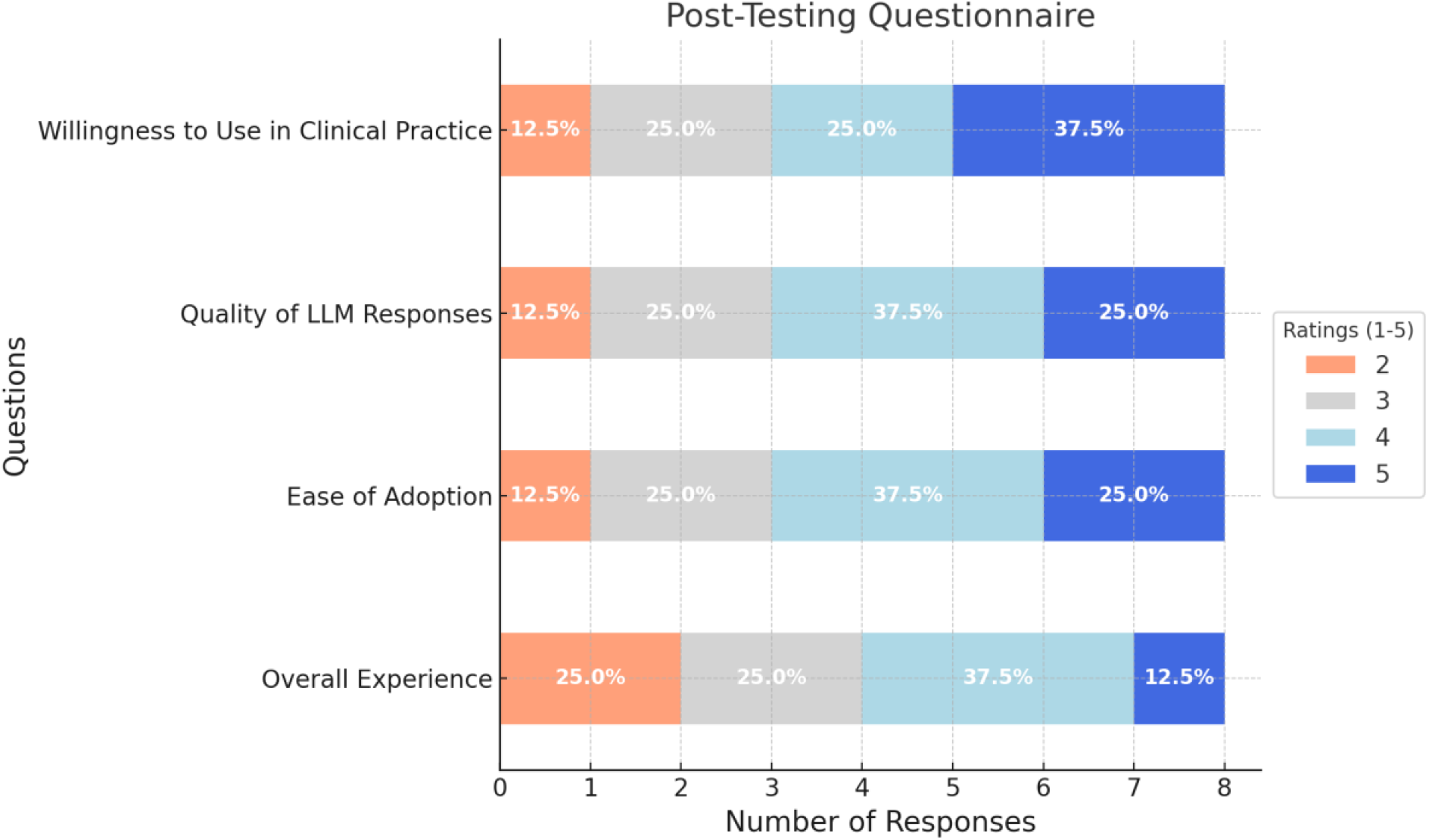
Post-testing questionnaire results (1: very low, 5: very high).

### Qualitative Findings

Several challenges in human-LLM interaction related to both human errors and technical limitations were observed (Table 3). Misleading or inaccurate search results attributable to human errors included inaccurate descriptions of image findings (e.g. describing the location of a cerebral cavernous venous malformation as “extra-axial”) or an omission of relevant imaging features (e.g. omission of multinodular morphology of a multinodular and vacuolating neuronal tumor; MVNT). LLM search, in contrast, exhibited bias based on clinical information irrelevant to the diagnosis (e.g. history of kidney transplantation in a patient with posterior reversible encephalopathy syndrome; PRES) or connotative terminology (e.g. the term “juxtacortical” was strongly associated with multiple sclerosis). Participant feedback on LLM-assisted differential diagnosis is illustrated in Table 4. Prior to the usability sessions, several participants expressed concerns about excessive reliance on the LLM system and consecutive impairment of their own radiological training. Following the testing, participants pointed out the ability to give flexible instructions regarding scope (e.g. quantity of differential diagnoses) and format (e.g. bullet points, table, sample images) of the search result as a key advantage over conventional internet search. Participants believed that usability of the LLM system for differential diagnosis could be enhanced by enabling voice-based interactions and improving the accuracy of returned sample images.

**Table 3:** Challenges during human-LLM interaction derived from observations and participant comments.

| Challenges | Examples |
| --- | --- |
| Omission of relevant clinical information | - Omitting the age in a patient with spinocerebellar ataxia (resulted in misleading suggestions such as atypical Parkinson syndrome) |
| Inclusion of clinical information irrelevant to the diagnosis | - Indicating recent history of head trauma in a patient with an incidental finding (benign enhancing foramen magnum lesion)<br>- Indicating a history of kidney transplantation in a patient with posterior reversible encephalopathy syndrome (PRES) |
| Inaccurate description of imaging findings | - Describing the location of a cerebral cavernous venous malformation as “subarachnoid” or “extra-axial”<br>- Searching for differential diagnoses of global brain atrophy in a patient with focal atrophy of the caudate head |
| Omission of relevant imaging features | - Omitting the bubbly or multinodular appearance of a multinodular and vacuolating neuronal tumor (MVNT) resulted in highly unspecific suggestions |
| Usage of connotative terminology | - Describing the location of a lesion as “juxtacortical” in a patient with MVNT (was strongly associated with multiple sclerosis) |
| LLM returning completely unrelated content (very few) | - Upon a request for sample images of progressive multifocal leukoencephalopathy (PML), images of the prime minister of India were returned |

**Table 4:** Participant comments on LLM-assisted differential diagnosis.

| Theme | Comments |
| --- | --- |
| Concerns<br>(pre-testing) | Residents might develop a strong dependence on the tool, educational effect could be impaired |
|  | Excessive reliance on LLM tool can promote intellectual laziness and carelessness in image interpretation |
|  | LLMs are known to create hallucinations, source of information is not always clear |
|  | The role of the radiologist might become obsolete as core tasks are increasingly overtaken by AI tools including LLMs |
| Benefits | Possibility to phrase open questions rather than rigid keyword searches is liberating |
|  | Links to information sources are embedded in the response |
|  | Instructions can be given regarding the scope (e.g. quantity of differential diagnoses suggested) and format of the response (e.g. table, bullet points) |
|  | The context of prior queries is retained so that follow-up questions can be posed |
|  | Useful to obtain a quick overview of a broad range of differential diagnoses |
| Suggestions for improvement | Interaction with the LLM system via voice |
|  | Improved search of relevant sample images |
|  | Quantitative data regarding the probability of certain diagnoses in the presence of specific imaging features |

## Discussion

In this study, we conducted usability experiments to explore the role of human-LLM collaboration in brain MRI differential diagnosis. Our results suggest that an LLM-assisted workflow has the potential to increase diagnostic accuracy as compared to conventional internet search. Yet, no clear effect on interpretation times and reader confidence was observed. Participant ratings of the LLM-assisted workflow were only neutral to slightly positive. These findings are consistent with the observed frustration and delays caused by misleading LLM outputs. These were frequently due to irrelevant or omitted information in prompts, highlighting the critical role of human readers to contextualize and filter available information. In addition, effective differential diagnosis was contingent on critical evaluation and validation of initial LLM recommendations. Readers typically conducted further research to correlate image findings with sample images of suggested diagnoses. Focused internet search on trusted websites (e.g. Radiopaedia) proved more effective for this purpose, indicating that the joint use of LLMs and conventional internet search might outperform the exclusive use of LLMs. A conceptual model illustrating the role of the human agent in LLM-assisted differential diagnosis and potential sources of error is shown in Figure 6. Altogether, our results underline the necessity to study collaborative efforts between humans and LLMs over LLMs in isolation to better reflect real-world conditions. While LLMs can augment human capabilities, traditional neuroradiological expertise remains indispensable for their effective utilization.

**Figure 6:**
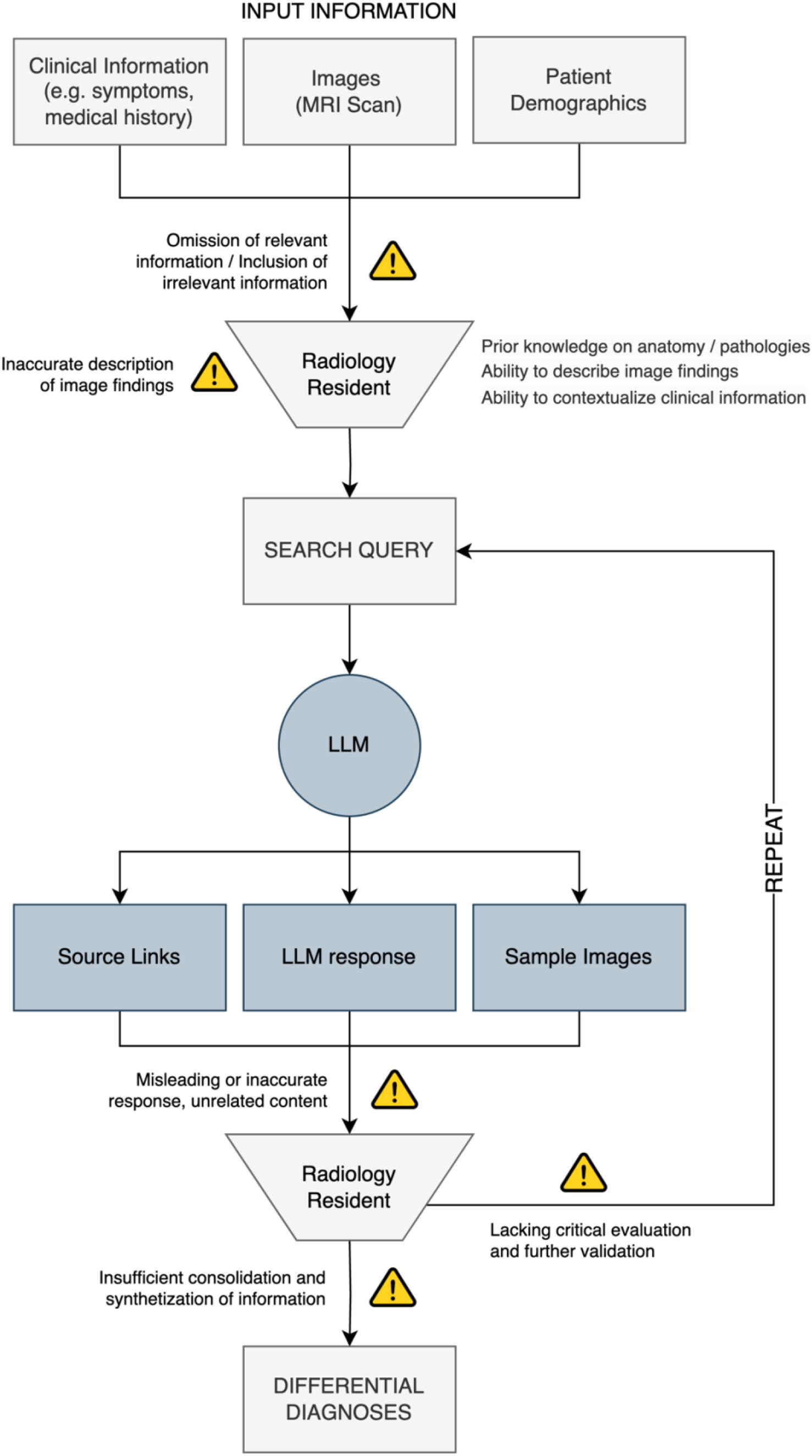
Conceptual model of human-LLM-Interaction for radiological differential diagnosis. Warning icons indicate potential sources of error in human-LLM collaboration.

Notably, rapid advancements in LLM technology demand continuous evaluations. Recent innovations, such as voice assistants enabling conversational interactions (e.g. as featured in OpenAI’s ChatGPT), are transforming LLM usability. Earlier, the introduction of speech recognition dictation systems has enhanced productivity in radiology departments (12). It is yet to be determined whether voice interaction with LLMs can yield similar benefits. The emergence of multimodal LLMs capable of processing image inputs is expected to create new possibilities. Few recent studies have demonstrated their potential to generate chest x-ray reports (13), and answer USMLE questions involving radiological images (14). In radiological differential diagnosis, direct image processing by LLMs could significantly alter human-LLM interactions by eliminating the need for human image descriptions. Seamless integration of LLMs into established health information systems could further boost productivity. For instance, prompt generation could be streamlined by directly importing clinical information from a RIS or selecting a key image from a PACS system as input.

This study has several limitations. First, only radiology residents with less than six months of experience in neuroradiology were included, based on the assumption that this group would be most likely to require additional research for differential diagnosis. It is unclear whether our findings would apply to more experienced readers. Second, human-LLM interactions were evaluated in a controlled environment but not in a real-world clinical setting, where frequent interruptions and intense workload might alter reader behavior. Third, the participants’ familiarity with the LLM system was limited, compared to their extensive experience with conventional internet search. Additional exposure and training could help users formulate more effective prompts, thereby reducing frustration and inefficiencies. Finally, a general-purpose LLM (GPT-4) was used. Models specifically trained for medicine (e.g. MED-PALM, GlassAI) (15–17) or radiology (18–20) are anticipated to further enhance clinical utility.

In conclusion, our study shows that human-LLM collaboration has the potential to enhance differential diagnosis of brain MRI but recognizes the necessity to mitigate both human and technical errors to maximize effectiveness.

## Data Availability

All data produced in the present study are available upon reasonable request to the authors

## Abbreviations

MRI: Magnetic Resonance Imaging
LLM: Large Language Model

## Notes

### Competing Interest Statement

The authors have declared no competing interest.

### Funding Statement

This study did not receive any funding

### Author Declarations

Ethics committee of the Technical University of Munich (School of Medicine and Health) waived ethical approval for this work.

